# Predictors of all-cause mortality among patients hospitalized with influenza, respiratory syncytial virus, or SARS-CoV-2

**DOI:** 10.1101/2022.03.31.22273111

**Authors:** Mackenzie A. Hamilton, Ying Liu, Andrew Calzavara, Maria E. Sundaram, Mohamed Djebli, Dariya Darvin, Stefan Baral, Rafal Kustra, Jeffrey C. Kwong, Sharmistha Mishra

## Abstract

**Background:** Shared and divergent predictors of clinical severity across respiratory viruses may support clinical and community responses in the context of a novel respiratory pathogen.

**Methods:** We conducted a retrospective cohort study to identify predictors of 30-day all-cause mortality following hospitalization with influenza (N=45,749; 2011-09 to 2019-05), respiratory syncytial virus (RSV; N=24,345; 2011-09 to 2019-04), or severe acute respiratory syndrome coronavirus 2 (SARS-CoV-2; N=8,988; 2020-03 to 2020-12; pre-vaccine) using population-based health administrative data from Ontario, Canada. Multivariable modified Poisson regression was used to assess associations between potential predictors and mortality. We compared the direction, magnitude, and confidence intervals of risk ratios to identify shared and divergent predictors of mortality.

**Results:** 3,186 (7.0%), 697 (2.9%) and 1,880 (20.9%) patients died within 30 days of hospital admission with influenza, RSV, and SARS-CoV-2, respectively. Shared predictors of increased mortality included: older age, male sex, residence in a long-term care home, and chronic kidney disease. Positive associations between age and mortality were largest for patients with SARS-CoV-2. Few comorbidities were associated with mortality among patients with SARS-CoV-2 as compared to those with influenza or RSV.

**Conclusions:** Our findings may help identify patients at greatest risk of illness secondary to a respiratory virus, anticipate hospital resource needs, and prioritize local prevention and therapeutic strategies to communities with higher prevalence of risk factors.

## BACKGROUND

The COVID-19 pandemic has put tremendous strain on hospital systems, and exposed long-standing issues in healthcare capacity.^1^ Knowing who is at highest risk of severe disease from respiratory viruses may support proactive clinical decision-making, and help distribute resources to healthcare settings with high prevalence of risk factors.^2^ This is particularly useful in the context of a new and emerging respiratory virus where information and resources are scarce.^2,3^

Several studies have compared shared and divergent predictors of severe disease among patients with influenza and respiratory syncytial virus (RSV)^4–9^, two respiratory viruses with high seasonal prevalence prior to the emergence of severe acute respiratory syndrome coronavirus 2 (SARS-CoV-2). However, few papers have compared predictors of severity across influenza, RSV, and SARS-CoV-2.

Communities are returning to pre-pandemic contact and exposure patterns, which may increase the risk of all respiratory infections. At the same time, laboratory diagnostic testing is transitioning to pre-pandemic approaches, where only a subset of hospitalized patients with viral respiratory or influenza-like illness receive laboratory-confirmed diagnoses.^10^ Thus, during periods of respiratory viral epidemics (particularly with novel emerging pathogens), shared predictors of severity across the most important respiratory viruses may: 1) reduce morbidity and mortality by prioritizing preventions (e.g. vaccinations), testing, and access to therapeutics (e.g. antivirals) and; 2) prepare healthcare settings that will require greater resources based on the prevalence of the underlying predictors.

We conducted an observational study using extensive health administrative data from Ontario, Canada to identify the direction and magnitude of shared and divergent predictors of 30-day all-cause mortality following hospitalization with influenza, RSV, or SARS-CoV-2 (prior to vaccine availability or variant emergence).

## METHODS

### Study setting and design

We conducted a retrospective cohort study of patients hospitalized with influenza, RSV or SARS-CoV-2 using population-based laboratory and health administrative data from Ontario, Canada (population 14.7 million^11^). Ontario’s healthcare system provides publicly funded physician services, laboratory testing, and hospital care for all residents with a provincial health card. Datasets used in this study were linked using unique encoded identifiers and analyzed at ICES^12^.

### Case definitions and outcomes

#### Hospitalization

We generated three study cohorts to assess predictors of severe outcomes among patients hospitalized with influenza, RSV, and SARS-CoV-2 respectively. Patients with influenza and RSV were identified using hospitalization data from the Canadian Institute for Health Information’s Discharge Abstract Database (DAD) during the 2010-11 to 2018-19 respiratory virus seasons. DAD captures administrative, clinical, and demographic information on all hospital discharges in Canada. Patients were considered hospitalized with influenza if their discharge abstract contained any of the following ICD-10 codes: J09, J10.0, J10.1, J10.8, J11.0, J11.1, or J11.8. Patients were considered hospitalized with RSV if their discharge abstract contained any of the following ICD-10 codes: J12.1, J20.5, J21.0, or B97.4. Case definitions were validated in Ontario against laboratory confirmation, and showed high specificity (influenza: 98%; RSV: 99%), and positive predictive values (influenza: 91%; RSV: 91%).^13^

We used DAD, the Ontario Laboratories Information System (OLIS), and the Public Health Case and Contact Management System (CCM) to identify patients hospitalized with SARS-CoV-2 between March 1 and December 1, 2020. OLIS is an electronic repository of Ontario’s laboratory test results, containing information on laboratory orders, patient demographics, provider information, and test results. CCM is a central data repository for all COVID-19 case management, contact management, and reporting in Ontario. Patients were considered hospitalized with SARS-CoV-2 if: 1) they were documented as hospitalized in DAD and had a positive polymerase chain reaction test for SARS-CoV-2 within 14 days before or 3 days after hospital admission; or 2) they were documented as hospitalized in CCM.

#### Mortality

Our primary outcome of interest was 30-day all-cause mortality following hospital admission with influenza, RSV or SARS-CoV-2. We used the Registered Persons Database (RPDB) and CCM to identify patients who died within 30 days of hospital admission. RPDB contains basic demographic information including age, sex, postal code, and date of death among all residents with an Ontario health card.

### Inclusion and exclusion criteria

Hospitalized patients were excluded if: they were not eligible for the Ontario Health Insurance Plan; their birthdate, sex, or postal code was missing from RPDB; their residential postal code was outside of Ontario; they were older than 105 years according to their birthdate in RPDB; or their recorded death date predated hospital admission (**Figure 1**). Only one hospitalization per patient was included (per season for influenza and RSV, and overall for SARS-CoV-2). Among patients hospitalized with influenza or RSV, we included the first hospital admission of the season. Among patients hospitalized with SARS-CoV-2, we included any hospitalization that resulted in death within 30 days of admission, or the first admission if no other admission was associated with 30-day mortality. Variation in inclusion criteria were due to suspected differences in hospital admission and discharge behavior across virus cohorts. For example, early in the pandemic, evidence suggested that patients hospitalized with SARS-CoV-2 had a relatively high likelihood of readmission within 60 days of discharge.^14^ Patients hospitalized with influenza or RSV were excluded if they were hospitalized outside of the respective respiratory virus season. Respiratory virus seasonality was defined as November to May for influenza, and November to April for RSV to align with case definitions from Hamilton et al.^13^, and to create the most inclusive time frame to capture seasonal virus activity in Ontario.^15^

**Figure 1.**
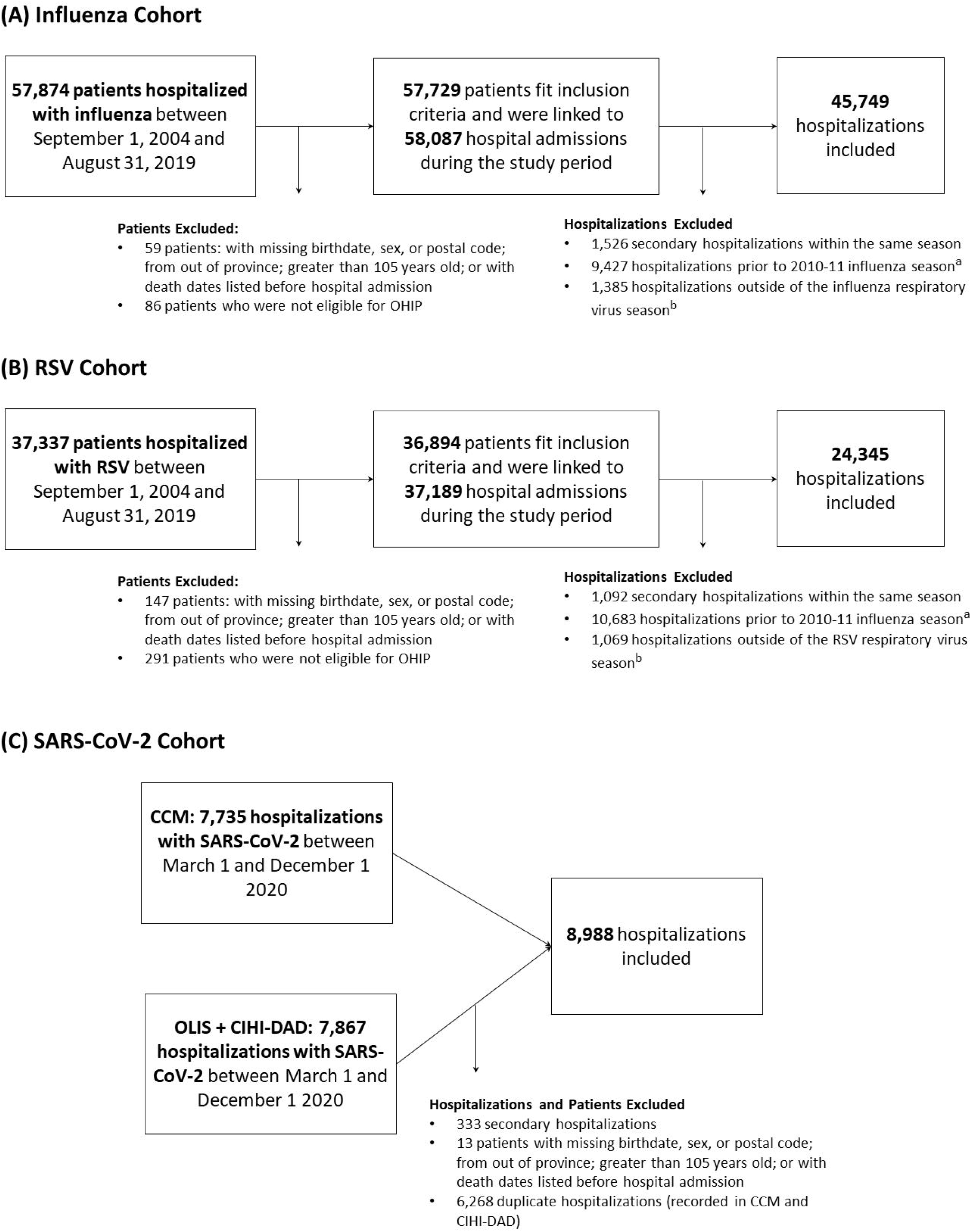
Study cohorts and exclusions. A) Influenza hospitalization cohort. B) RSV hospitalization cohort. C) SARS-CoV-2 hospitalization cohort. Exclusions were made in the order in which they appear. RSV, respiratory syncytial virus; SARS-CoV-2, severe acute respiratory syndrome coronavirus 2; OHIP, Ontario health insurance plan. †Influenza and RSV hospitalizations prior to the 2010-11 respiratory virus season were excluded to reduce selection bias due to changes in testing behavior following the H1N1 influenza epidemic in 2009-10. ‡Virus seasonality was defined as November through May, and November through April for influenza and RSV, respectively.

### Predictors of 30-day all-cause mortality

We selected potential predictors of 30-day all-cause mortality *a priori*. Variables were considered if they had documented or suspected associations with respiratory virus acquisition or severity, or healthcare access in peer-reviewed, published literature.

#### Demographic Characteristics

We used data from RPDB to describe pertinent individual-level demographic characteristics including age, sex, and residence in rural neighbourhoods.^16^ Rural neighbourhoods were defined as those outside commuting zones of population centres (i.e. centres with more than 10,000 residents).

We used aggregated 2016 Canadian census data to describe neighbourhood-level social determinants of health associated with risk of respiratory virus acquisition^17–19^, access to care, and discrimination within health care settings^20,21^ including: income^22^, household size^23^, and “ethnic concentration”^24^ (herein referred to as percent racialized). Neighbourhood-level variables were categorized into quintiles (i.e. 1 = 20% of neighbourhoods with lowest values; 5 = 20% of neighbourhoods with highest values). Patients were assigned a quintile according to their residential postal code. We describe derivation of the neighbourhood-level determinants of health in depth in **Supplementary text**.

#### Underlying Health Conditions

Pertinent underlying health conditions included: asthma, chronic obstructive pulmonary disease (COPD), hypertension, cardiac ischemic disease, congestive heart failure, stroke, dementia or frailty, chronic kidney disease, advanced liver disease, and immunosuppression (i.e. patients with a cancer diagnosis in the past 5 years, human immunodeficiency virus, solid organ or bone marrow transplant, or another immunodeficiency condition).^25,26^ We used validated case definitions and health administrative data to classify each individual-level health condition. Case definitions and validity are described in detail in **Supplementary Table 1**.

#### Other Covariates

Other predictors of severe outcomes included residence in a long-term care home (LTCH)^27,28^, and seasonal immunization against influenza^29–31^. We used the Chronic Care Reporting System, pharmacist billing claims in the Ontario Drug Benefits Database (ODB), and physician billing claims in the Ontario Health Insurance Plan (OHIP) database to determine whether individuals resided in a LTCH. We used ODB and OHIP to identify patients vaccinated against influenza between October 1 of the season of hospital admission and 14 days prior to admission. Relevant vaccination claim codes and drug identification numbers are outlined in **Supplementary Table 2**.

### Statistical analyses

Data processing and analyses were conducted using SAS version 9.4 (SAS Institute, Cary, NC). Frequencies and proportions were used to describe the distribution of risk factors among individuals hospitalized with influenza, RSV, or SARS-CoV-2. Modified Poisson regression (i.e. Poisson regression with a robust error variance) was used to assess the association between predictors and 30-day all-cause mortality. Modified Poisson regression was used over logistic regression to estimate risk ratios and avoid misinterpretation of odds ratios obtained from logistic regression.^32^ We calculated unadjusted and adjusted relative risk of dying within 30 days of hospital admission per predictor among each hospitalization cohort. Adjusted models included all other predictors. We qualitatively compared the direction, magnitude, and 95% confidence interval of associations among respective cohorts of hospitalized patients to identify shared and divergent predictors of 30-day all-cause mortality.

## RESULTS

### Characteristics of patients hospitalized with influenza, RSV or SARS-CoV-2

We observed 45,749 influenza hospitalizations, 24,345 RSV hospitalizations, and 8,988 SARS-CoV-2 hospitalizations after applying inclusion and exclusion criteria (**Figure 1**). Patients hospitalized with RSV were younger than patients hospitalized with influenza or SARS-CoV-2 (median age RSV patients = 1 year; median age influenza patients = 71 years; median age SARS-CoV-2 patients = 70 years). Only 47% of RSV patients presented with at least one comorbidity as compared to 84% of influenza patients and 82% of SARS-CoV-2 patients. **Table 1** compares additional characteristics of hospitalized patients by virus and 30-day all-cause mortality.

**Table 1.** Descriptive characteristics of patients hospitalized with influenza, respiratory syncytial virus, or SARS-CoV-2.

| Characteristic | Influenza |  | RSV |  | SARS-CoV-2 |  |
| --- | --- | --- | --- | --- | --- | --- |
|  | n (%) | Deaths (%) | n (%) | Deaths (%) | n (%) | Deaths (%) |
| **Total** | 45,749 | 3,186 | 24,345 | 697 | 8,988 | 1,880 |
| **Demographics** |  |  |  |  |  |  |
| Age group |  |  |  |  |  |  |
| 0-4 | 4,863 (10.6) | 11 (0.3) | 17,157 (70.5) | 20 (2.9) | 65 (0.7) | 0 (0) |
| 5-19 | 2,363 (5.2) | 12 (0.4) | 557 (2.3) | 7 (1.0) | 67 (0.8) | 0 (0) |
| 20-49 | 4,466 (9.8) | 95 (3.0) | 421 (1.7) | 19 (2.7) | 1,367 (15.2) | 36 (1.9) |
| 50-64 | 6,519 (14.2) | 305 (9.6) | 969 (4.0) | 59 (8.5) | 2,150 (23.9) | 186 (9.9) |
| 65-74 | 7,030 (15.4) | 466 (14.6) | 1,298 (5.3) | 88 (12.6) | 1,674 (18.6) | 347 (18.5) |
| 75-84 | 9,981 (21.8) | 827 (26.0) | 1,837 (7.5) | 190 (27.3) | 1,826 (20.3) | 535 (28.5) |
| ≥85 | 10,527 (23.0) | 1,470 (46.1) | 2,106 (8.7) | 314 (45.1) | 1,839 (20.5) | 776 (41.3) |
| Male sex | 21,831 (47.7) | 1,551 (48.7) | 12,603 (51.8) | 302 (43.3) | 4,768 (53.1) | 1,029 (54.7) |
| Living in rural area | 3,818 (8.3) | 260 (8.2) | 2,243 (9.2) | 41 (5.9) | 253 (2.8) | 53 (2.8) |
| Long-term care resident | 3,048 (6.7) | 814 (25.5) | 721 (3.0) | 168 (24.1) | 1,355 (15.1) | 620 (33.0) |
| Immunized against seasonal influenza | 14,718 (32.2) | 1,083 (34.0) | 4,106 (16.9) | 287 (41.2) | 3,077 (34.2) | 694 (36.9) |
| **Underlying Health Conditions** |  |  |  |  |  |  |
| Asthma | 12,838 (28.1) | 762 (23.9) | 6,630 (27.2) | 190 (27.3) | 1,777 (19.8) | 345 (18.4) |
| COPD | 11,924 (26.1) | 1,183 (37.1) | 2,593 (10.7) | 273 (39.2) | 1,098 (12.2) | 342 (18.2) |
| Cardiac ischemic disease | 9,301 (20.3) | 1,044 (32.8) | 1,717 (7.1) | 225 (32.3) | 1,063 (11.8) | 341 (18.1) |
| Congestive heart failure | 12,240 (26.8) | 1,507 (47.3) | 2,769 (11.4) | 349 (50.1) | 1,609 (17.9) | 540 (28.7) |
| Hypertension | 28,811 (63.0) | 2,644 (83.0) | 5,379 (22.1) | 580 (83.2) | 5,912 (65.8) | 1,590 (84.6) |
| Diabetes | 16,036 (35.1) | 1,373 (43.1) | 2,828 (11.6) | 295 (42.3) | 3,795 (42.2) | 996 (53.0) |
| Dementia/frailty | 11,149 (24.4) | 1,557 (48.9) | 2,451 (10.1) | 368 (52.8) | 2,604 (29.0) | 940 (50.0) |
| Stroke | 3,999 (8.7) | 445 (14.0) | 738 (3.0) | 102 (14.6) | 679 (7.6) | 224 (11.9) |
| Chronic kidney disease | 9,729 (21.3) | 1,091 (34.2) | 2,007 (8.2) | 250 (35.9) | 2,120 (23.6) | 711 (37.8) |
| Immunosuppression | 7,222 (15.8) | 578 (18.1) | 1,936 (8.0) | 174 (25.0) | 602 (6.7) | 146 (7.8) |
| Advanced liver disease | 1,305 (2.9) | 115 (3.6) | 246 (1.0) | 26 (3.7) | 252 (2.8) | 47 (2.5) |
| **Neighbourhood-level social determinants of health<sup>g</sup>** |  |  |  |  |  |  |
| Income quintile |  |  |  |  |  |  |
| Missing | 149 (0.3) | 11 (0.3) | 147 (0.6) | 1-5<sup>†</sup> | 67 (0.8) | 10 (0.5) |
| 1 (lowest income) | 12,199 (26.7) | 773 (24.3) | 5,967 (24.5) | 202 (29.0) | 2,617 (29.1) | 541 (28.8) |
| 2 | 10,187 (22.3) | 798 (25.0) | 5,162 (21.2) | 164 (23.5) | 2,081 (23.2) | 483 (25.7) |
| 3 | 8,802 (19.2) | 597 (18.7) | 4,754 (19.5) | 113 (16.2) | 1,823 (20.3) | 404 (21.5) |
| 4 | 7,564 (16.5) | 482 (15.1) | 4,527 (18.6) | 110 (15.8) | 1,293 (14.4) | 234 (12.5) |
| 5 (highest income) | 6,848 (15.0) | 525 (16.5) | 3,788 (15.6) | 103-107<sup>†</sup> | 1,107 (12.3) | 208 (11.1) |
| Household size quintile |  |  |  |  |  |  |
| Missing | 416 (0.9) | 30 (0.9) | 380 (1.6) | 10 (1.4) | 139 (1.6) | 34 (1.8) |
| 1 (smallest household size) | 11,947 (26.1) | 911 (28.6) | 4,614 (19.0) | 202 (29.0) | 1,729 (19.2) | 410 (21.8) |
| 2 | 8,319 (18.2) | 636 (20.0) | 4,507 (18.5) | 129 (18.5) | 1,168 (13.0) | 253 (13.5) |
| 3 | 6,173 (13.5) | 445 (14.0) | 3,512 (14.4) | 105 (15.1) | 905 (10.1) | 205 (10.9) |
| 4 | 9,605 (21.0) | 665 (20.9) | 5,821 (23.9) | 155 (22.2) | 2,156 (24.0) | 457 (24.3) |
| 5 (largest household size) | 9,289 (20.3) | 499 (15.7) | 5,511 (22.6) | 96 (13.8) | 2,891 (32.2) | 521 (27.7) |
| Percent racialized quintile |  |  |  |  |  |  |
| Missing | 420 (0.9) | 27 (0.8) | 328 (1.3) | 7 (1.0) | 98 (1.1) | 19 (1.0) |
| 1 (least percent racialized) | 7,035 (15.4) | 512 (16.1) | 3,573 (14.7) | 85 (12.2) | 579 (6.4) | 115 (6.1) |
| 2 | 7,482 (16.4) | 540 (16.9) | 4,124 (16.9) | 99 (14.2) | 843 (9.4) | 212 (11.3) |
| 3 | 8,179 (17.9) | 601 (18.9) | 4,510 (18.5) | 135 (19.4) | 1,355 (15.1) | 340 (18.1) |
| 4 | 9,578 (20.9) | 743 (23.3) | 5,039 (20.7) | 174 (25.0) | 1,914 (21.3) | 380 (20.2) |
| 5 (most percent<br>racialized) | 13,055 (28.5) | 763 (23.9) | 6,771 (27.8) | 197 (28.3) | 4,199 (46.7) | 814 (43.3) |

RSV, respiratory syncytial virus; SARS-CoV-2, severe acute respiratory syndrome coronavirus 2; COPD, chronic
obstructive pulmonary disease.
†Cells with less than five patients have been suppressed to prevent individual identification.

### Common predictors of 30-day all-cause mortality

Patients hospitalized with SARS-CoV-2 had the highest crude 30-day all-cause mortality rate (SARS-CoV-2 crude mortality rate = 20.9%; influenza crude mortality rate = 7.0%; RSV crude mortality rate = 2.9%).

In unadjusted models, shared predictors of mortality included: older age, residence in a LTCH, immunization against seasonal influenza, COPD, cardiac ischemic disease, congestive heart failure, hypertension, diabetes, dementia/frailty, stroke, and chronic kidney disease (**Figure 2, Supplementary Table 3**). Larger magnitudes of association between older age and mortality were observed among patients hospitalized with SARS-CoV-2 [unadjusted relative risk (RR) among 85+ versus 50-64 = 4.88; 95% confidence interval (CI) = 4.16 to 5.72] versus influenza (unadjusted RR among 85+ versus 50-64 = 2.99; 95% CI = 2.65 to 3.37) or RSV (unadjusted RR among 85+ versus 50-64 = 2.53; 95% CI = 1.93 to 3.32). All other shared predictors of mortality showed larger magnitudes of association among patients hospitalized with RSV (**Figure 2**).

**Figure 2.**
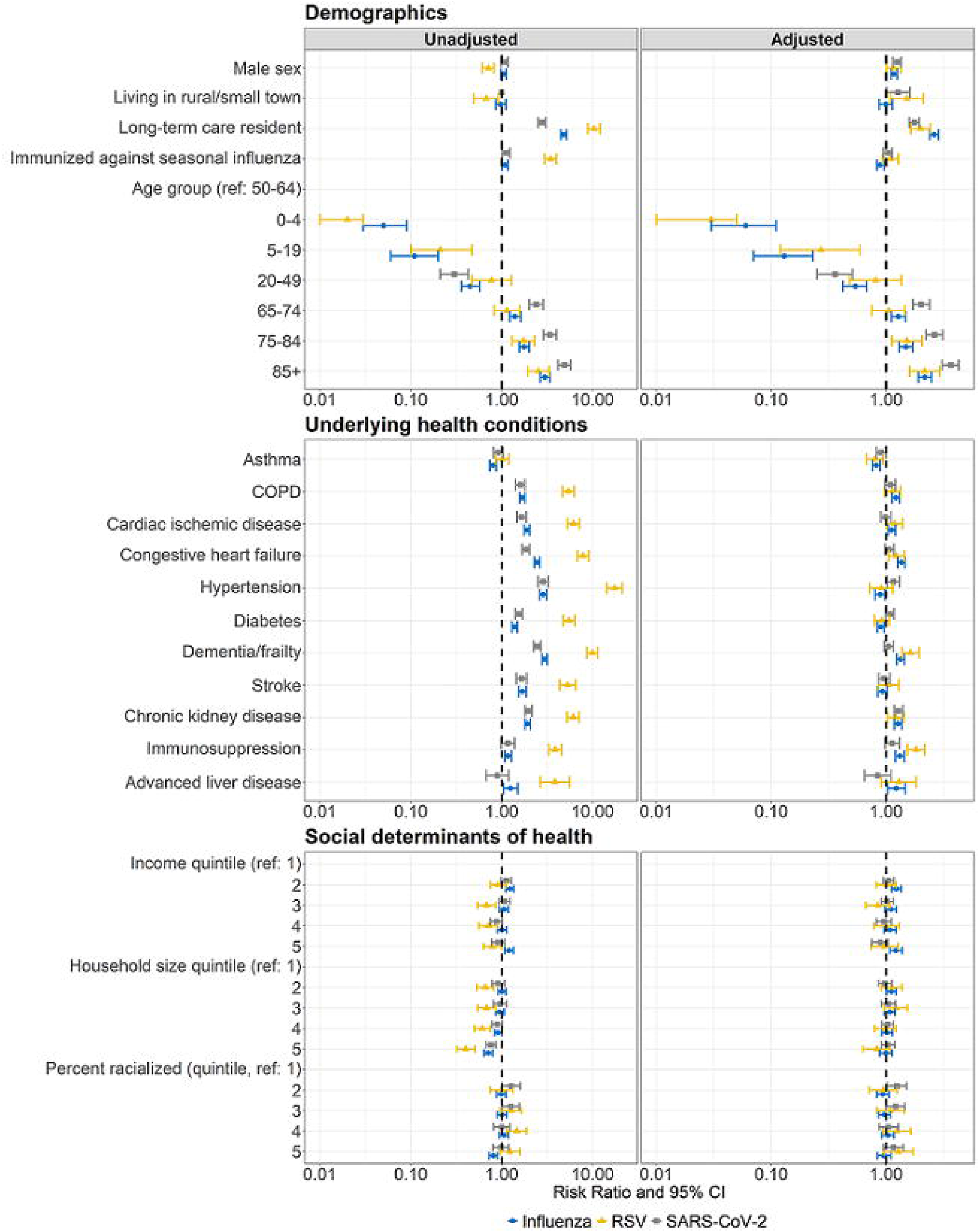
Unadjusted and adjusted predictors of 30-day all-cause mortality among patients hospitalized with influenza, RSV or SARS-CoV-2. Modified Poisson regression was used to calculate associations between predictors and 30-day all-cause mortality. Adjusted models included all predictors. Influenza and RSV adjusted models additionally included season of hospital admission. Associations are presented as risk ratios (points) and 95% confidence intervals (error bars). RSV, respiratory syncytial virus; SARS-CoV-2, severe acute respiratory syndrome coronavirus 2; COPD, chronic obstructive pulmonary disease; CI, confidence interval.

In adjusted models, shared predictors of mortality included: older age, male sex, residence in a LTCH, and chronic kidney disease (**Figure 2, Supplementary Table 4**). Similar to unadjusted models, we observed larger magnitudes of association between older age and 30-day all-cause mortality among patients hospitalized with SARS-CoV-2 [SARS-CoV-2: adjusted RR (95% CI) 85+ versus 50-64 = 3.64 (3.08 to 4.30); influenza: adjusted RR (95% CI) 85+ versus 50-64 = 2.18 (1.91 to 2.48); RSV: adjusted RR (95% CI) 85+ versus 50-64 = 2.16 (1.60 to 2.92)]. The magnitude and direction of associations between male sex, residence in a LTCH, and chronic kidney disease, and mortality were similar among patients hospitalized with all three viruses.

### Notable differences among predictors of 30-day all-cause mortality

Rural residence was associated with increased 30-day all-cause mortality among patients hospitalized with RSV (adjusted RR = 1.52, 95% CI = 1.09 to 2.12) and SARS-CoV-2 (adjusted RR = 1.27, 95% CI = 1.01 to 1.61), but not among patients hospitalized with influenza (adjusted RR = 1.00, 95% CI = 0.87 to 1.14). Immunization against seasonal influenza was associated with decreased 30-day all-cause mortality among patients hospitalized with influenza (adjusted RR = 0.89, 95% CI = 0.83 to 0.96) but not patients hospitalized with RSV (adjusted RR = 1.09, 95% CI = 0.93 to 1.28) or SARS-CoV-2 (adjusted RR = 1.04, 95% CI = 0.95 to 1.13). Finally, cardiac ischemic disease, congestive heart failure, dementia/frailty, and immunosuppression were associated with increased all-cause mortality among patients hospitalized with influenza and RSV after adjustment for all other predictors, but not among patients hospitalized with SARS-CoV-2.

## DISCUSSION

We identified shared and divergent predictors of mortality among patients hospitalized with influenza, RSV, or SARS-CoV-2 using population-based health administrative data from Ontario, Canada. In multivariable models, common predictors of 30-day all-cause mortality following hospitalization included older age, male sex, residence in a LTCH, and chronic kidney disease.

Older age and male sex were predictive of increased mortality across all respiratory virus cohorts, which aligns with numerous studies from high-income countries^4–9,33–35^, and confirms the need to consider age and sex in clinical practice. The magnitude of association between older age and mortality was largest among patients with SARS-CoV-2, confirming robust evidence that age is an important predictor of severity among COVID-19 patients, and should be used to guide targeted COVID-19 preventions and therapeutics.^35,36^

Residence in a LTCH was also a common predictor of 30-day all-cause mortality; however, associations were weaker among patients hospitalized with SARS-CoV-2. Differences in magnitudes of association may be due to greater selection bias of LTCH residents hospitalized with SARS-CoV-2 in comparison to those with influenza or RSV. For example, in Ontario, less than one quarter of COVID-19-positive LTCH residents were hospitalized prior to death, compared to nearly 80% of COVID-19-positive community residents during the first wave of the pandemic ^37^. It has been suggested that LTCH residents with SARS-CoV-2 may have been less likely to be hospitalized due to advanced care directives and/or informal policies that discouraged transfers of critically ill residents^37–39^. The difference in hospitalizations prior to death narrowed in the pre-vaccination second wave of the pandemic in Ontario^37^ suggesting that the selection biases may have been specific to wave 1, and may not be reflective of past influenza or RSV seasonal epidemics.

Similar to previous studies^40^, chronic kidney disease increased risk of 30-day all-cause mortality with similar magnitudes of effect among patients hospitalized with influenza, RSV or SARS-CoV-2. Several other comorbidities were important predictors of mortality among patients with influenza or RSV, but not SARS-CoV-2 despite their known associations with SARS-CoV-2 severity.^26^ These comorbidities may have been associated with mortality among influenza and RSV patients, but not SARS-CoV-2 patients due to: 1) smaller sample size of the latter; 2) greater hospitalization rates of less severe patients with comorbidities who were infected with SARS-CoV-2 (i.e. selective hospitalization of patients with comorbidities due to limited understanding of the virus and disease trajectory); and/or 3) true clinical differences between patients requiring hospitalization with SARS-CoV-2 versus seasonal influenza or RSV.^41–43^ Moreover, age may act as an effect measure modifier on the relationship between comorbidities and mortality due to SARS-CoV-2. More research is needed to compare the immunological and clinical disease progression of influenza, RSV, and SARS-CoV-2 to better explain observed differences in risk by comorbidity.

We did not observe associations between area-level social determinants of health and 30-day all-cause mortality following hospitalization with all three viruses, despite their associations with infection transmission risk.^17–21^ Lack of associations may be due to misclassification of neighbourhood-level social determinants of health (as these metrics were derived from the 2016 census), ecological fallacy, or adjustment of mediators in the causal pathway between income, household size, or racialization, and 30-day all-cause mortality.

This study is limited by potential misclassification of influenza and RSV cases, as we identified patients using their hospitalization discharge codes rather than diagnostic test results. However, case definitions were validated against a population of hospitalized patients who received diagnostic testing for influenza or RSV in the Ontario population.^13^ The case definitions had high specificity (influenza = 98%; RSV = 99%) and positive predictive values (influenza = 91%; RSV = 91%). Thus, misclassification of influenza and RSV hospitalization is likely rare. Moreover, the use of influenza and RSV case definitions allowed us to obtain hospitalization data across more respiratory virus seasons, increasing the generalizability of our findings.

Moreover, the study outcome as defined may capture deaths attributable to the virus or deaths in the context of an incidental infection (i.e., death *with* the virus). The additional public health dataset available for SARS-CoV-2 (i.e., CCM) allowed us to compare death outcomes of hospitalized patients with SARS-CoV-2 (this dataset documents whether cause of death was due to, or likely due to COVID-19) to their mortality within 30 days of hospitalization in health administrative data (i.e., RPDB). 30-day all-cause mortality had 97% positive predictive value against death due to COVID-19 in CCM. Thus, we expect 3% of outcomes to potentially reflect deaths with SARS-CoV-2. In the absence of approaches to adequately address this misclassification for all viruses, we acknowledge the small bias as an important limitation.

This study is also limited by a lack of data on other important predictors of respiratory infection severity such as pregnancy^44,45^, obesity^46^, and individual-level social determinants (e.g. economic marginalization and racialization), which are known to mediate quality of hospitalized care and rates of respiratory virus infection^17–21^. When using our results to inform prioritization of services, or to develop clinical prediction tools, we must consider these limitations so that other at-risk patients do not fall through the cracks.

Finally, to provide insights on shared predictors of mortality in the context of a novel, emerging pathogen, we purposefully restricted the study period of SARS-CoV-2 to exclude hospitalizations of patients vaccinated against SARS-CoV-2, or those with SARS-CoV-2 variants. Future work would benefit from comparisons of predictors of mortality among patients hospitalized with influenza, RSV or SARS-CoV-2 variants and/or breakthrough infections in the context of a vaccine to better inform responses to re-emerging respiratory pathogens.

Our results add to the growing literature base comparing similarities and differences in clinical disease progression of patients hospitalized with influenza and SARS-CoV-2^41–43^, and have three important implications for clinical care and health systems. First, shared predictors of mortality could be used to identify, target, and prioritize hospitalized patients who are at greatest risk of death for prevention (e.g. vaccines), testing (e.g. rapid tests) and therapeutics (e.g. antivirals) in the context of a novel respiratory pathogen. Second, the underlying prevalence of shared predictors in a given geography could help prepare health systems for, and efficiently allocate health resources during, emergence of a novel respiratory pathogen. Finally, differences in observed predictors of mortality across the three viruses signal the importance of sufficient virus-specific laboratory testing to ensure at-risk individuals are not left behind.

## Conclusion

We identified common predictors of 30-day all-cause mortality following hospitalization with SARS-CoV-2, influenza, or RSV in a population-based cohort from Ontario, Canada. Shared predictors of mortality may help identify patients at greatest risk for syndromic clinical management of illness from respiratory viruses, anticipate local resource needs (e.g. for communities and hospitals), and prioritize prevention and therapeutic strategies during rapid emergence of respiratory viruses.

## Data Availability

The dataset from this study is held securely in coded form at ICES. While legal data sharing agreements between ICES and data providers (e.g., healthcare organizations and government) prohibit ICES from making the dataset publicly available, access may be granted to those who meet pre-specified criteria for confidential access, available at www.ices.on.ca/DAS. The full dataset creation plan and underlying analytic code are available from the authors upon request, understanding that the computer programs may rely upon coding templates or macros that are unique to ICES and are therefore either inaccessible or may require modification.

## Acknowledgments

We thank IQVIA Solutions Canada Inc. for use of their Drug Information File. Parts of this material are based on data and/or information compiled and provided by the Canadian Institute for Health Information (CIHI) and by Ontario Health (OH). However, the analyses, conclusions, opinions and statements expressed herein are solely those of the authors, and do not reflect those of the data sources; no endorsement by CIHI or OH is intended or should be inferred. JCK is supported by a Clinician-Scientist Award from the University of Toronto Department of Family and Community Medicine. SM is supported by a Tier 2 Canada Research Chair in Mathematical Modeling and Program Science.

## AUTHOR CONTRIBUTION STATEMENT

MAH, MD, MS, RK, JCK and SM were involved in conceptualization, design and methodological decisions of the study. JCK and SM led supervision, administration and funding acquisition for the study. Supervision was supported by RK and SB. YL curated and cleaned data for analyses of patients hospitalized with influenza and RSV supported by AC, and AC curated and cleaned data of patients hospitalized with SARS-CoV-2. MAH conducted the formal analysis and investigation of predictors of mortality among patients hospitalized with influenza and RSV, supported by MD. MS conducted the formal analysis and investigation of predictors of mortality among patients hospitalized with SARS-CoV-2. DD led the visualization of analyses supported by MAH. MAH drafted the original manuscript and led the review and editing process. All authors edited and/or reviewed, and approved the final manuscript.

