## Supplementary Material for "Predictors of all-cause mortality among patients hospitalized with influenza, respiratory syncytial virus, or SARS-CoV-2"

**SUPPLEMENTARY TEXT**

**Derivation of neighbourhood-level social determinants of health**

We analyzed associations between area-level social determinants of health and 30-day all-cause mortality among individuals hospitalized with influenza, respiratory syncytial virus (RSV) or COVID-19. Three social determinants of health with known associations to access of health care and risk of respiratory virus acquisition were analyzed, including: income, household size, and percent racialized. We describe the data sources and derivation of area-level variables below:

1) *Income quintile:*

To derive neighbourhood income quintiles, we used data from the 2016 Canadian census aggregated at the dissemination area (DA) level – the smallest geographic unit at which census data are collected.^1^ DAs were ranked into quintiles according to their total 2016 income (before tax) per person equivalent^2^ in relation to other DAs in the same census metropolitan area or census agglomeration unit (i.e. DAs in quintile 1 are among 20% of DAs with the lowest income in their city; DAs in quintile 5 are among 20% of DAS with the highest income in their city). Quintiles were estimated per census metropolitan area or census agglomeration unit to adjust for variability in cost of living across Ontario. If a DA was missing median income, neighbouring DAs were used to estimate median income and calculate the income per person equivalent. Patients were assigned neighbourhood income quintiles according to their postal codes in the registered person’s database (RPDB).

2) *Household size quintile:*

To derive average household size quintiles, we used data from the 2016 Canadian census aggregated by DA. DAs were ranked into quintiles according to the average number of people per dwelling per DA (i.e. quintile 1= 20% of Ontario DAs with the least people per dwelling; quintile 5 = 20% of Ontario DAs with the most people per dwelling).^3^ Patients were assigned household size quintiles according to their postal codes in the RPDB. Quintiles 1, 2, 3, 4, and 5 included DA’s with an average household size between: 0 to 2.1; 2.2 to 2.4; 2.5 to 2.6; 2.7 to 3.0; and 3.1 to 5.7 persons per dwelling respectively.

3) *Percent racialized quintile:*

We used Public Health Ontario’s ethnic concentration quintile^4^ to categorize neighbourhoods by their concentration of individuals who are racialized. This dimension categorizes neighbourhoods according to the proportion of the population who may experience marginalization related to racism or discrimination. Derivation of Ontario’s ethnic concentration quintiles are described elsewhere^4^. In brief, an ethnic concentration principle component score was derived from two neighbourhood-level census variables: 1) the proportion of recent immigrants^5,6^; and 2) the proportion of people who self-identify as ‘visible minorities.’^7^ Neighbourhoods were ranked into quintiles according to the principle component score. Patients were assigned neighbourhood ethnic concentration quintiles according to their postal codes in the RPDB.

**SUPPLEMENTARY TABLES**

**Supplementary Table 1.** Definitions of underlying medical conditions among hospitalized patients.

| **Medical condition** | **Data sources** | **Definition** | **Validated** | **Validity metrics** |
| --- | --- | --- | --- | --- |
| **Advanced liver disease** | OHIP,  DAD,  NACRS,  SDS | **Defined as patients with cirrhosis or decompensated cirrhosis.**  **Patients with cirrhosis**, defined as those with:  2 or more physician visits for cirrhosis (in OHIP) or  1 or more hospitalization for cirrhosis (in DAD, NACRS, SDS)  According to:   - OHIP diagnostic code 571 - ICD-9 diagnostic codes 456.1, 571.2, or 571.5 - ICD-10 diagnostic codes I85.9, I98.2, K70.3,K71.7, or K74.6   **OR**  **Patients with decompensated cirrhosis**, defined as those with:  1 or more physician visits classified as cirrhosis (in OHIP, diagnostic code 571) and  1 or more hospitalizations for decompensated cirrhosis (in DAD, NACRS, SDS) according to:   - ICD-9 diagnostic codes 456.0, 456.2, 572.2, 572.3, 572.4, 782.4, or 789.5 - ICD-10 diagnostic codes I85.0, I86.4, I98.20, I98.3, K721, K729, K76.6, K76.7, R17, or R18 - CCI codes 1.NA.13.BA-FA, 1.NA.13.BA-X7, 1.NA.13.BA-BD, 1.KQ.76GP-NR, or 1.OT.52.HA - CCP codes 1006 or 6691 - OHIP codes J057 or Z591 | Yes^8^ | **Cirrhosis:**  Sensitivity = 95%  Specificity = 78%  **Decompensated Cirrhosis:**  Sensitivity = 99%  Specificity = 79% |
| **Asthma** | DAD,  SDS,  OHIP | 1 or more hospital admissions (DAD, SDS) for asthma, or  2 or more physician visits (OHIP) for asthma within 2 years  According to:   - ICD-9 diagnostic code 493 - ICD-10 diagnostic codes J45 or J46 - OHIP diagnostic code 493 | Yes^9–11^ | Sensitivity = 80.6%  Specificity = 81.4%  PPV = 72.5%  NPV = 87.3% |
| **Cancer** | Ontario Cancer Registry | Patients in Ontario Cancer Registry with any cancer diagnosed in the 5 years prior to cohort start date, except for non-melanoma skin cancer (ICD-O-3 Topography = C44 and Morphology = 87xx3)  The Ontario Cancer Registry identifies patients with cancer according to 4 major data sources:   - Hospitalization admission and same day surgery data (DAD, SDS, NACRS) - Pathology reports with any mention of cancer - Records of patients referred to Cancer Care Ontario’s eight regional cancer centers or the Princess Margaret Hospital – the specialized institutions treating cancer patients in Ontario - Death certificates with cancer recorded as the underlying cause of death | No | N/A |
| **Cardiac ischemic disease** | DAD,  SDS | 1 or more hospital admissions for cardiac ischemic disease (DAD) in the past 5 years (i.e. angina, chronic ischemic heart disease, or myocardial infarction), or  1 or more procedures related to cardiac ischemic disease (DAD, SDS) in the past 20 years (i.e. coronary artery bypass grafting or percutaneous coronary interventions)  According to:   - ICD-9 diagnostic codes: 413, 4140, 4148, 4149, 410, 411, 412 - ICD-10 diagnostic codes: I20, I21, I22 or I25 - CCI procedure codes: 1IJ50, 1IJ5, 1IJ57 or 1IJ76 - CCP procedure codes 481, 4802 or 4803 | No | N/A |
| **Chronic kidney disease** | DAD,  NACRS, OHIP | 1 or more hospitalizations for chronic kidney disease (in DAD or NACRS) in the past 5 years or  1 or more physician claims for treatment of chronic kidney disease (in OHIP) in the past 5 years or  1 or more dialysis codes in each of the 3 months prior to the cohort start date  According to:   - ICD-10 diagnostic codes E102, E112, E132, E142, I12, I13, N08, N18, or N19 - OHIP diagnostic codes 403 or 585   **OR**  **Patients on chronic dialysis** in the year before cohort start date, defined as those with at least 2 of any of the following codes in OHIP, DAD or SDS separated by at least 90 days, but less than 150 days:   - OHIP service codes R849, G323, G325, G326, G860, G862, G865 G863, G866, G330, G331, G332, G333, G861, G082, G083, G085, G090, G091, G092, G093, G094, G095, G096, G294, G295, G864, H540, or H740 - CCI procedure codes 5195, or 6698 - CCP procedure code 1PZ21 | Yes^12,13^ | **Chronic kidney disease:**  Sensitivity = 32.7%  Specificity = 96.9%  PPV = 65.4%  NPV = 88.8%  **Chronic dialysis:**  Sensitivity = 78.63%  Specificity = 99.96%  PPV = 80.65%  NPV = 99.96% |
| **Chronic obstructive pulmonary disease** | DAD,  SDS,  OHIP | 1 or more hospital admission (DAD or SDS) for COPD or  1 or more physician visit (OHIP) for COPD  According to:   - ICD-9 diagnostic codes 491, 492 or 496; - ICD-10 diagnostic codes J41, J42, J43 or J44 - OHIP diagnostic codes 491, 492 or 496 | Yes^14^ | Sensitivity: 85.0%  Specificity: 78.4% |
| **Congestive heart failure** | DAD,  NACRS,  SDS,  OHIP | 1 or more hospital admissions (DAD, SDS) for congestive heart failure among individuals aged 40 years or older, or  1 or more physician claim (OHIP) or emergency department record (NACRS) for congestive heart failure within one year, followed by a second record with a congestive heart failure diagnosis from any source among individuals aged 40 years or older  According to:   - ICD-9 diagnosis code: 428 - ICD-10 diagnosis codes: I500, I501, or I509 - OHIP diagnosis code: 428 | Yes^15^ | Sensitivity: 84.8%  Specificity: 97.0%  PPV: 55.6% |
| **Dementia or frailty** | DAD,  SDS,  OHIP,  ODB | **Patients with dementia**, defined as those with:  3 physician visits for dementia (in OHIP), each recorded at least 30 days apart in a 2-year period or  1 hospitalization or same day surgery for dementia (in DAD, SDS) or  1 ODB claim for a dispensed dementia medication (cholinesterase inhibitors)  According to:   - OHIP diagnostic codes 290 or 331 - ICD-9 diagnostic codes 0461, 290.0, 290.1, 290.2, 290.3, 290.4, 294, 331.0, 331.1 or 331.5 - ICD-10 diagnostic codes F00, F01, F02, F03 or G30   **OR**   - **Frail patients,** defined as those with a Hospital Frailty Risk Score greater than 15 according to Gilbert et al. algorithm ^16^ and DAD data in the 5 years prior to cohort start date. | Yes^16–18^ | **Dementia**  Sensitivity: 79.3%  Specificity 99.1%  PPV 80.4%  NPV 99.0% |
| **Diabetes (non-gestational)** | DAD,  SDS,  OHIP | **Children/youth (<19):**  4 or more physician visits (OHIP) for diabetes (diagnosis code 250) within 2 years or  1 physician visit (OHIP) claimed with a fee code: Q040, K029, K030, K045 and K046  **Adults 19+:**  2 or more physician visits (in OHIP) for diabetes within 1 year, or  1 hospital admission for diabetes (DAD or SDS) or  1 ODB drug claim for a diabetes medication  According to:   - ICD-9 diagnostic code 250 - ICD-10 diagnostic codes E10, E11, E13, E14 | Yes^19–21^ | **Children/youth (<19):**  Sensitivity: 82.8%  Specificity: 98.9%  **Adults (19+):**  Sensitivity: 90.0%  Specificity: 97.7%  PPV: 82.6% |
| **Hypertension** | DAD,  SDS,  OHIP | 1 or more hospital admission (DAD, SDS) for hypertension, or  1 physician claim (OHIP) for hypertension, followed by a second OHIP claim or hospital admission for hypertension within 2 years from the first  According to:   - ICD-9 diagnosis codes: 401x, 402x, 403x, 404x, or 405x - ICD-19 diagnosis codes: I10, I11, I12, I13 or I15 - OHIP diagnosis codes: 401, 402, 403, 404 or 405.   Note: we excluded individuals with gestational hypertension defined as those with hypertension records between 120 days before and 180 days after a gestational admission date. | Yes^22^ | Sensitivity: 72%  Specificity: 95%  PPV: 87% |
| **Immunosuppressed** | OHIP,  Canadian Organ Replacement Register,  DAD | **Patients with HIV**, defined as:  3 or more physician claims (OHIP) for HIV within 3 years according to OHIP diagnosis codes: 042, 043 or 044  **OR**  **Patients who have received an organ transplant**, as documented in the Canadian Organ Replacement Register. The Canadian Organ Replacement Register is a national information system which records the activity and outcomes of vital organ transplantation.  **OR**  **Patients who have received an allogenic/autologous bone marrow transplant,** defined as:  1 or more physician claims (OHIP) for an allogenic or autologous bone marrow transplant according to:   - CCP code: 53.0 - CCI codes: 1WY19, 1LZ19HHU7, or 1L19HHU8 - OHIP fee code: Z426   **OR**  **Patients with an immunodeficient condition** (i.e. sickle-cell disease; hereditary immunodeficiency; neutropenia; functional disorders of polymorphonuclear neutrophils and anomalies of leukocytes; hyposplenism, hypersplenism and chronic congestive splenomegaly; asplenia) defined as:  1 or more hospitalization (DAD) for an immunodeficient condition  According to:   - ICD-9 diagnosis codes: 282.6, 279, 288.0, 288.1, 288.2, 289.4, 289.5, or 759.0 - ICD-10 diagnosis codes: D57.0 to D57.2, D57.8, D80 to D84, D89.8, D89.9, D70, D71 to D72, D73.0 D73.1, D73.2 or Q89.0 | **HIV:** Yes^23^  **Bone marrow transplant**: No  **Immunodeficient condition**: No | **HIV:**  Sensitivity: 96.2%  Specificity: 99.6%  **Bone marrow transplant**: N/A  **Immunodeficient condition**: N/A |
| **Stroke** | DAD,  NACRS, | **Patients who have had a transient ischemic attack**, defined as:  1 or more hospitalization (DAD), or emergency department visit (NACRS) for a transient ischemic attack according to  According to:   - ICD-9 diagnosis codes: 435, 3623 - ICD-10 diagnosis codes: G450, G451, G452, G453, G458, G459, H340   **OR**  **Patients who have had an acute ischemic stroke**, defined as:  1 or more hospitalizations (DAD) with a main diagnosis of acute ischemic stroke  According to:   - ICD-9 diagnosis codes: 43301, 43311, 43321, 43331, 43391, 434, 436 - ICD-10 diagnosis codes: I63, I64, H34.1 | No | N/A |

OHIP, Ontario health insurance plan database; DAD, Canadian Institute for Health information Discharge Abstract Database; SDS, Canadian Institute for Health Information Same-Day Surgery Database; NACRS, Canadian Institute for Health Information National Ambulatory Care Reporting System; ICD-9, International Classification of Disease Revision 9; ICD-10, International Classification of Disease Revision 10; CCI, Canadian Classification of Health Interventions; CCP, Canadian Classification of Diagnostic, Therapeutic, and Surgical Procedures; ODB, Ontario Drug Benefits Database; HIV, human immunodeficiency virus.

**Supplementary Table 2.** Databases and claim codes to identify patients immunized against seasonal influenza.

| **Database** | **Code / DIN** | **Code Description/Product Name** |
| --- | --- | --- |
| Ontario Health Insurance Plan Database | G590 | Influenza agent |
|  | G591 | Injection of an influenza agent – sole reason |
|  | G592 | Administration of intranasal influenza vaccine |
|  | Q130 | Influenza tracking code |
|  | Q590 | Basic flu shot fee-per-visit premium FHN/FHO |
|  | Q690 | Influenza agent – with visit, each inject – N.P |
|  | Q691 | Influenza agent sole reason – N.P. |
| Ontario Drug Benefit Claims Database | 02015986 | FLUVIRAL |
|  | 02223929 | VAXIGRIP |
|  | 02269562 | INFLUVAC |
|  | 02346850 | AGRIFLU |
|  | 02362384 | FLUAD |
|  | 02365936 | FLUZONE |
|  | 02420643 | FLUZONE QUADRIVALENT |
|  | 02420686 | FLUVIRAL |
|  | 02420783 | FLULAVAL TETRA |
|  | 02426544 | FLUMIST QUADRIVALENT |
|  | 02428881 | AGRIFLU |
|  | 02432730 | FLUZONE QUADRIVALENT |
|  | 09857501 | FLUZONE |

**Supplementary Table 3.** Unadjusted predictors of 30-day all-cause mortality among patients hospitalized with influenza, respiratory syncytial virus, or SARS-CoV-2 (2020-03 to 2020-12).

| **Risk Factor** | **Risk ratio (95% CI)** | | |
| --- | --- | --- | --- |
|  | **Influenza** | **RSV** | **SARS-CoV-2** |
| **Demographics** | | | |
| Age group (ref: 50-64) |  |  |  |
| 0-4 | 0.05 (0.03-0.09) | 0.02 (0.01-0.03) | NA |
| 5-19 | 0.11 (0.06-0.20) | 0.21 (0.10-0.47) | NA |
| 20-49 | 0.45 (0.36-0.57) | 0.77 (0.47-1.28) | 0.30 (0.21 – 0.43) |
| 65-74 | 1.40 (1.22-1.62) | 1.14 (0.82-1.57) | 2.40 (2.01 – 2.86) |
| 75-84 | **1.77 (1.56-2.01)** | **1.73 (1.30-2.30)** | **3.39 (2.87 – 4.00)** |
| ≥85 | **2.99 (2.65-3.37)** | **2.53 (1.93-3.32)** | **4.88 (4.16 – 5.72)** |
| Male sex | 1.05 (0.98-1.12) | 0.71 (0.61-0.82) | 1.07 (0.98 – 1.17) |
| Living in rural/small town^a^ | 0.97 (0.86-1.11) | 0.67 (0.49-0.92) | 1.00 (0.76 – 1.32) |
| Long-term care resident | **4.80 (4.47-5.16)** | **10.3 (8.82-12.1)** | **2.77 (2.52 – 3.05)** |
| Immunized against seasonal influenza | **1.09 (1.01-1.17)** | **3.44 (2.97-3.99)** | **1.12 (1.02 – 1.23)** |
| **Underlying health conditions** | | | |
| Asthma | 0.80 (0.74-0.87) | 1.02 (0.86-1.20) | 0.91 (0.81 – 1.03) |
| COPD | **1.68 (1.57-1.80)** | **5.39 (4.65-6.25)** | **1.60 (1.42 – 1.80)** |
| Cardiac ischemic disease | **1.90 (1.77-2.04)** | **6.16 (5.29-7.17)** | **1.65 (1.47 – 1.86)** |
| Congestive heart failure^b^ | **2.45 (2.29-2.62)** | **7.82 (6.77-9.03)** | **1.85 (1.67 – 2.04)** |
| Hypertension | **2.85 (2.60-3.12)** | **17.4 (14.3-21.2)** | **2.85 (2.52 – 3.23)** |
| Diabetes | **1.39 (1.30-1.49)** | **5.50 (4.75-6.36)** | **1.54 (1.41 – 1.69)** |
| Dementia or frailty^c^ | **2.96 (2.77-3.16)** | **9.91 (8.59-11.4)** | **2.45 (2.24 – 2.68)** |
| Stroke^d^ | **1.69 (1.53-1.86)** | **5.29 (4.33-6.47)** | **1.66 (1.44 – 1.90)** |
| Chronic kidney disease^e^ | **1.93 (1.80-2.07)** | **6.10 (5.26-7.09)** | **1.97 (1.80 – 2.16)** |
| Immunosuppression^f^ | 1.17 (1.08-1.28) | 3.86 (3.27-4.55) | 1.17 (0.99 – 1.39) |
| Advanced liver disease | 1.25 (1.05-1.50) | 3.84 (2.65-5.57) | 0.89 (0.67 – 1.19) |
| **Neighbourhood social determinants of health^g^** | | | |
| Income quintile (ref: 1^st^ quintile)^h^ |  |  |  |
| 2 | 1.23 (1.11-1.35) | 0.91 (0.75-1.12) | 1.12 (0.99 – 1.27) |
| 3 | 1.06 (0.95-1.18) | 0.68 (0.54-0.85) | 1.07 (0.94 – 1.22) |
| 4 | 1.01 (0.90-1.13) | 0.71 (0.56-0.89) | 0.88 (0.75 – 1.02) |
| 5 (highest income per person) | 1.21 (1.08-1.34) | 0.79 (0.63-1.00) | 0.91 (0.77 – 1.07) |
| Household size quintile (ref: 1^st^ quintile)^i^ |  |  |  |
| 2 | 1.01 (0.91-1.11) | 0.66 (0.53-0.81) | 0.91 (0.78 – 1.07) |
| 3 | 0.95 (0.86-1.06) | 0.68 (0.54-0.86) | 0.96 (0.81 – 1.13) |
| 4 | 0.91 (0.83-1.00) | 0.61 (0.50-0.75) | 0.89 (0.78 – 1.02) |
| 5 (largest household size) | **0.71 (0.64-0.79)** | **0.40 (0.32-0.51)** | **0.76 (0.67 – 0.86)** |
| Percent racialized (quintile, ref: 1^st^ quintile)^j^ |  |  |  |
| 2 | 0.99 (0.88-1.11) | 1.00 (0.75-1.33) | 1.27 (1.01 – 1.59) |
| 3 | 1.01 (0.90-1.13) | 1.26 (0.96-1.64) | 1.26 (1.02 – 1.56) |
| 4 | 1.06 (0.95-1.18) | 1.46 (1.13-1.88) | 1.00 (0.81 – 1.23) |
| 5 (greatest percent racialized) | 0.81 (0.73-0.90) | 1.23 (0.96-1.58) | 0.98 (0.80 – 1.19) |

Modified poisson regression was used to calculate associations between predictors and 30-day all-cause mortality. Associations are presented as risk ratios and 95% confidence intervals. Values in bold indicate significant 95% confidence intervals across all three hospitalization cohorts. RSV, respiratory syncytial virus; SARS-CoV-2, severe acute respiratory syndrome coronavirus 2; COPD, chronic obstructive pulmonary disease.

^a^Defined as a city or town with a population of less than 10,000.

^b^Among patients aged 40 years or older.

^c^Defined as a Hospital Frailty Risk Score^16^ greater than 15.

^d^Defined as history of a transient ischemic attack or an acute ischemic stroke.

^e^Within the past 5 years.

^f^Defined as patients with a cancer diagnosis in the past 5 years, who were HIV positive, who had an organ or bone marrow transplant, or who had another immunodeficiency condition.

^g^Social determinants of health were captured at the level of the Canadian census dissemination area.

^h^Income was measured as a per-person equivalent. Quintiles were generated per census agglomeration unit or census metropolitan area to adjust for variability in cost of living across the province. Quintile ranges vary across the province.

^i^Range household size quintile 1 = 0 - 2.1 persons per dwelling; quintile 2 = 2.2 – 2.4 persons per dwelling; quintile 3 = 2.5 – 2.6 persons per dwelling; quintile 4 = 2.7 – 3 persons per dwelling; quintile 5 = 3.1-5.7 persons per dwelling.

^j^As defined by the “ethnic concentration quintile” from the Ontario Marginalization Index.^4^ Quintiles were created by ranking dissemination areas on a principle component score; thus, quintile ranges are uninterpretable.

**Supplementary Table 4.** Adjusted predictors of 30-day all-cause mortality among patients hospitalized with influenza, respiratory syncytial virus, or SARS-CoV-2 (2020-03 to 2020-12).

| **Risk Factor** | **Risk ratio (95% CI)** | | |
| --- | --- | --- | --- |
|  | **Influenza** | **RSV** | **SARS-CoV-2** |
| **Demographics** | | | |
| Age group (ref: 50-64) |  |  |  |
| 0-4 | 0.06 (0.03-0.11) | 0.03 (0.01-0.05) | NA |
| 5-19 | 0.13 (0.07-0.23) | 0.27 (0.12-0.59) | NA |
| 20-49 | 0.54 (0.42-0.68) | 0.81 (0.48-1.36) | 0.36 (0.25 – 0.51) |
| 65-74 | 1.28 (1.11-1.48) | 1.05 (0.75-1.46) | 2.02 (1.71 – 2.40) |
| 75-84 | **1.49 (1.30-1.70)** | **1.52 (1.12-2.05)** | **2.64 (2.23 – 3.12)** |
| ≥85 | **2.18 (1.91-2.48)** | **2.16 (1.60-2.92)** | **3.64 (3.08 – 4.30)** |
| Male sex | **1.17 (1.10-1.25)** | **1.16 (1.01-1.35)** | **1.25 (1.15 – 1.35)** |
| Living in rural/small town^a^ | 1.00 (0.87-1.14) | 1.52 (1.09-2.12) | 1.27 (1.01 – 1.61) |
| Long-term care resident | **2.62 (2.40-2.86)** | **2.00 (1.64-2.43)** | **1.76 (1.59 – 1.94)** |
| Immunized against seasonal influenza | 0.89 (0.83-0.96) | 1.09 (0.93-1.28) | 1.04 (0.95 – 1.13) |
| **Underlying health conditions** | | | |
| Asthma | 0.82 (0.76-0.89) | 0.80 (0.68-0.94) | 0.90 (0.82 – 1.00) |
| COPD | 1.22 (1.13-1.31) | 1.14 (0.97-1.33) | 1.09 (0.98 – 1.21) |
| Cardiac ischemic disease | 1.12 (1.04-1.21) | 1.18 (1.01-1.39) | 0.99 (0.90 – 1.10) |
| Congestive heart failure^b^ | 1.36 (1.26-1.46) | 1.23 (1.05-1.44) | 1.07 (0.98 – 1.17) |
| Hypertension | 0.89 (0.80-0.98) | 0.91 (0.72-1.15) | 1.16 (1.02 – 1.31) |
| Diabetes | 0.90 (0.84-0.96) | 0.92 (0.79-1.07) | 1.08 (0.99 – 1.17) |
| Dementia or frailty^c^ | 1.34 (1.24-1.45) | 1.63 (1.38-1.94) | 1.05 (0.96 – 1.16) |
| Stroke^d^ | 0.93 (0.85-1.02) | 1.05 (0.86-1.29) | 0.96 (0.86 – 1.08) |
| Chronic kidney disease^e^ | **1.27 (1.18-1.37)** | **1.22 (1.03-1.43)** | **1.28 (1.18 – 1.40)** |
| Immunosuppression^f^ | 1.32 (1.21-1.44) | 1.82 (1.54-2.16) | 1.13 (0.98 – 1.31) |
| Advanced liver disease | 1.23 (1.03-1.47) | 1.29 (0.91-1.82) | 0.84 (0.65 – 1.10) |
| **Neighbourhood social determinants of health^g^** | | | |
| Income quintile (ref: 1^st^ quintile)^h^ |  |  |  |
| 2 | 1.23 (1.12-1.35) | 1.01 (0.82-1.23) | 1.05 (0.95 – 1.17) |
| 3 | 1.10 (0.98-1.23) | 0.85 (0.67-1.08) | 1.02 (0.91 – 1.16) |
| 4 | 1.08 (0.96-1.22) | 1.01 (0.78-1.30) | 0.96 (0.82 – 1.11) |
| 5 (highest income per person) | 1.22 (1.08-1.38) | 0.97 (0.74-1.27) | 0.89 (0.75 – 1.04) |
| Household size quintile (ref: 1^st^ quintile)^i^ |  |  |  |
| 2 | 1.11 (1.01-1.22) | 1.12 (0.91-1.38) | 0.98 (0.86 – 1.12) |
| 3 | 1.08 (0.96-1.20) | 1.22 (0.96-1.54) | 1.05 (0.91 – 1.21) |
| 4 | 1.02 (0.92-1.14) | 0.98 (0.79-1.22) | 1.03 (0.92 – 1.16) |
| 5 (largest household size) | 1.00 (0.88-1.13) | 0.83 (0.63-1.09) | 1.04 (0.91 – 1.19) |
| Percent racialized (quintile, ref: 1^st^ quintile)^j^ |  |  |  |
| 2 | 0.94 (0.83-1.06) | 0.94 (0.71-1.25) | 1.24 (1.02 – 1.51) |
| 3 | 0.97 (0.86-1.09) | 1.09 (0.83-1.44) | 1.21 (1.00 – 1.46) |
| 4 | 1.04 (0.92-1.17) | 1.24 (0.94-1.64) | 1.05 (0.87 – 1.28) |
| 5 (greatest percent racialized) | 0.96 (0.84-1.10) | 1.28 (0.96-1.72) | 1.16 (0.95 – 1.41) |

Modified poisson regression was used to calculate adjusted associations between predictors and 30-day all-cause mortality. Models were adjusted for all risk factors presented. Influenza and RSV models were additionally adjusted for season of hospital admission. Associations are presented as risk ratios and 95% confidence intervals. Values in bold indicate significant 95% confidence intervals across all three hospitalization cohorts. RSV, respiratory syncytial virus; SARS-CoV-2, severe acute respiratory virus syndrome coronavirus 2; COPD, chronic obstructive pulmonary disease.

^a^Defined as a city or town with a population of less than 10,000.

^b^Among patients aged 40 years or older.

^c^Defined as a Hospital Frailty Risk Score^16^ greater than 15.

^d^Defined as history of a transient ischemic attack or an acute ischemic stroke.

^e^Within the past 5 years.

^f^Defined as patients with a cancer diagnosis in the past 5 years, who were HIV positive, who had an organ or bone marrow transplant, or who had another immunodeficiency condition.

^g^Social determinants of health were captured at the level of the Canadian census dissemination area.

^h^Income was measured as a per-person equivalent. Quintiles were generated per census agglomeration unit or census metropolitan area to adjust for variability in cost of living across the province. Quintile ranges vary across the province.

^i^Range household size quintile 1 = 0 - 2.1 persons per dwelling; quintile 2 = 2.2 – 2.4 persons per dwelling; quintile 3 = 2.5 – 2.6 persons per dwelling; quintile 4 = 2.7 – 3 persons per dwelling; quintile 5 = 3.1-5.7 persons per dwelling.

^j^As defined by the “ethnic concentration quintile” from the Ontario Marginalization Index.^4^ Quintiles were created by ranking dissemination areas on a principle component score; thus, quintile ranges are uninterpretable.
